# Initial Heart Rate Score Predicts New Onset Atrial Fibrillation in Pacemaker Patients

**DOI:** 10.1101/2023.02.07.23285627

**Authors:** Katsuhide Hayashi, Haruhiko Abe, Brian Olshansky, Arjun D. Sharma, Paul W. Jones, Nickolas Wold, David Perschbacher, Ritsuko Kohno, Mark Richards, Bruce L. Wilkoff

## Abstract

**Background:** Heart Rate Score (HRSc), the percent of all atrial paced and sensed event in the largest 10 bpm rate histogram bin of a pacemaker, predicts survival in patients with cardiac devices. No correlation between HRSc and development of atrial fibrillation (AF) has been reported.

**Objective:** To evaluate the relationship between pacemaker post-implantation HRSc and newly-developed AF incidence.

**Methods:** Patients with dual-chamber pacemakers, implanted 2013-2017, with ALTITIUDE remote monitoring data with ≥600,000 beats of histogram data collected at baseline were included (N=34,543). HRSc was determined from the post-implantation histogram data during the initial 3 months. Patients were excluded if they had AF, defined as atrial high-rate episodes >5 minutes or >1% of right atrial beats >170 bpm during the initial 3-months post-implantation. New AF, after the baseline period, was defined by each of the following: >1%, >10% or >25% of atrial beats >170 bpm or Atrial Tachycardia Response (ATR) events >24 hr.

**Results:** Patients were followed a median of 2.8 (1.0-4.0) years. Patients with initial HRSc≥70% were older, had higher %RA pacing, had lower %RV pacing and were more likely programmed with rate-response vs subjects with HRSc<70%. The incidence of AF increased in proportion to HRSc (Log-Rank P-value <0.001); results were insensitive to AF definition. Initial HRSc (HR:1.07, 95% CI:1.05-1.09; P<0.0001) independently predicted AF after adjusting for age, gender, % RV pacing and rate-response programming. The %RA pacing and initial HRSc correlated.

**Conclusion:** HRSc predicts subsequent AF independent of well-known risk factors in pacemaker patients.

## Introduction

Heart rate score (HRSc), a novel parameter, is defined by the percentage of all paced and sensed atrial beats in the tallest 10-beat/minute heart rate histogram bin, recorded by a cardiac implantable pacemaker (PM) or implantable cardioverter-defibrillator (DDD-ICD) with an atrial lead, (Figure 1). A HRSc ≥70%, early post-implantation, correlates with mortality in patients without atrial fibrillation (AF) who have ICDs^1^ or cardiac resynchronization therapy defibrillators (CRT-Ds).^2,3^

**Figure 1.**
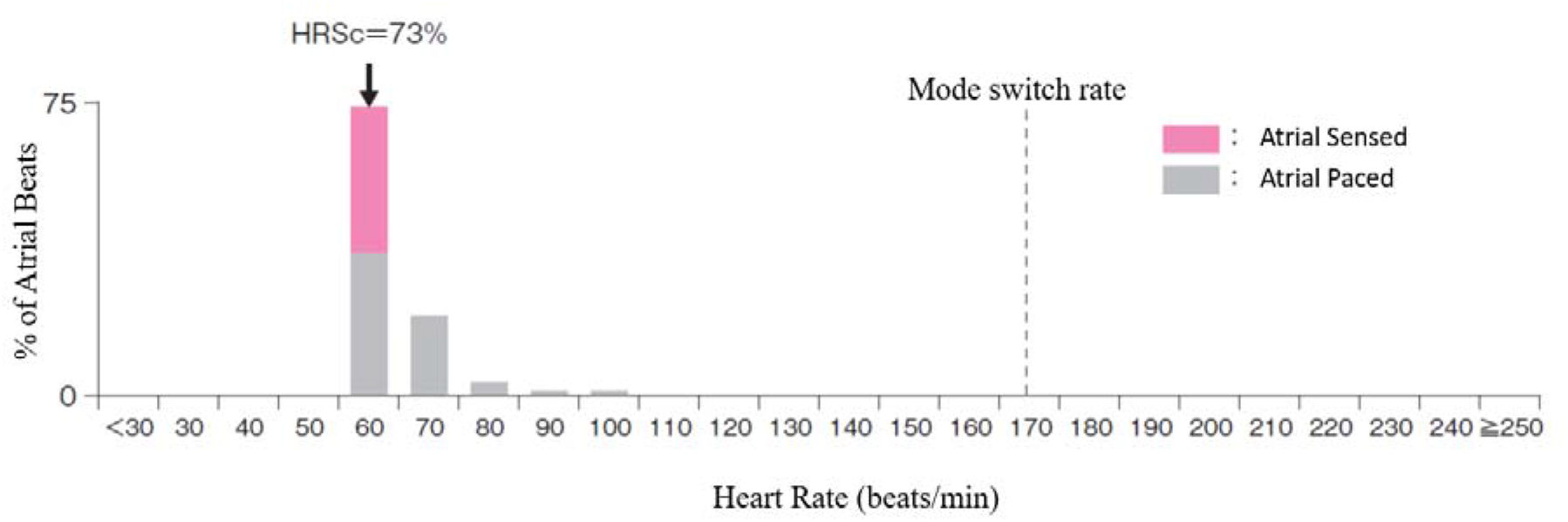
Example of HRSc. This figure shows an example of HRSc. Here, HRSc is calculated as 73%. HRSc indicates heart rate score.

Atrial High-Rate Episodes (AHREs), detected by cardiac devices, is associated with increased risk of stroke, systemic embolism, heart failure, and mortality.^4,5^ In the ASSERT trial, AHREs >190 beats/min for >6 minutes were associated with a significantly increased risk of ischemic stroke or systemic embolism. In sub-analysis of the ASSERT trial, AHREs >24 hours were associated with increased risk of subsequent stroke or systemic embolism (adjusted hazard ratio 3.24, 95% confidence interval [CI] 1.51-6.95).^6^ Unfortunately, no established data from electrocardiograms or device interrogation predicts development of AF.

In previous studies of HRSc, subjects with atrial tachyarrhythmias >24 hours detected by cardiac devices were excluded since frequent or persistent AF distorts the assessment of HRSc.^1,2^ Remotely monitored databases of PM patients offer the opportunity to assess associations between interrogated data from cardiac devices and clinical outcomes as recorded in device data.

The current retrospective analysis employs a large remote monitoring database and evaluates the relationship between the initial HRSc and subsequent AF in patients with dual-chamber PMs. No study has evaluated the relationship of HRSc with AF. We focused on the incidence of AF developing for the first time after determining a lack of AF during the baseline 3 month observation. We hypothesized that a higher initial HRSc in patients with a PM predicts subsequent occurrences of AF.

## Methods

### Study Cohort

This study, approved by the Boston Scientific Governance Board (Boston Scientific Company, Boston, USA), is a retrospective analysis of deidentified data from the LATITUDE remote monitoring cardiac implantable electronic device database. All subjects included underwent implantation of a dual-chamber Boston Scientific Corporation (Boston Scientific Corporation, Marlboro, MA) pacemaker from 2013-2017 in the United States and were regularly monitored remotely via the LATITUDE^®^ remote monitoring patient system.

Patients were excluded if they had:

1) Single chamber or biventricular PM

2) First LATITUDE transmission >3 months after implantation

3) Less than 600,000 beats to calculate HRSc

4) Device-determined AF detection during the first 3 months after implantation

5) Crossover to another programming mode in the first 3 months post-implant

6) Programming to non-atrial tracking mode (VVI, VVIR, DDI, DDIR) and/or lower pacing rate different than 60 bpm

7) Devices from centers opting out of data sharing

All patients with >1% of right atrial beats ≥170 beats/min or mode switch episodes ≥5 minutes were considered to have device-determined AF. Data were analyzed retrospectively. No individual subject was studied nor were patients identified; therefore, Institutional Review Board approval was not required.

### Heart Rate Score (HRSc) and Baseline Period

HRSc, used in all analyses, was determined from remote interrogation data from the dual chamber PM (Figure 1) during the baseline period (the first 3 months after PM implantation). The HRSc, calculated at the last LATITUDE transmission during the baseline period, was designated the initial HRSc. Only patients without device-determined AF in the baseline period and with >600,000 beats were included.

HRSc was analyzed as a continuous variable in 10 bpm increments and via three zones: <30%, 30%-69%, and ≥70%. These three zones were based on J-Statistic analysis of optimal cutpoint for prediction of outcome and are consistent with prior HRSc research.^2,3^

### Outcome

The endpoint was device-determined AF after the baseline period. The primary AF endpoint was >1% of atrial beats ≥170 beats/min. There were 4 secondary AF endpoints: 1) >5% of atrial beats ≥170 beats/min, 2) >10% of atrial beats ≥170 beats/min, 3) >25% of atrial beats ≥ 170 beats/min, 4) ATR mode switch episodes ≥24 hours.

The ATR algorithm continually monitors for sensed atrial events which are equal to or above the ATR (170 beats/min for 8 cycles: nominal value). Mode switch was designed to limit tracking of atrial arrhythmias by automatically switching to a non-tracking mode when programmed ATR criteria were met.

### Follow-up Data Analysis

Occurrence of AF was examined using stored LATITUDE remote monitoring data and tracked during follow-up periods. The relationship between the initial HRSc and occurrence of each device-determined AF definitions was assessed.

### Statistical Analysis

Continuous variables were expressed as mean±standard deviation (SD) or median (25^th^-75^th^), depending on the distribution of data, whereas categorical variables were expressed as counts and percentages. Demographics for the different HRSc groups were compared using χ^2^ tests for categorical variables and *F* tests for continuous variables. J-Statistic analysis was used to identify optimal cutpoint for prediction of outcome. Kaplan-Meier analyses evaluated freedom from AF after the baseline period for the three initial HRSc groups (<30%, 30-69%, and ≥70%). Log-rank tests were performed to compare group classified by initial HRSc. Multivariable Cox proportional hazard models were performed, adjusted for age, sex, cumulative percentage of ventricular pacing (% RV pacing), and rate-response programming. P<0.05 was considered statistically significant. All statistical analyses were performed using SAS Version 9.4 (SAS Institute, Cary, NC).

## Results

### Subjects Demographics

A total of 136,980 dual-chamber pacemakers were implanted between 2013-2017 in subjects followed with remote monitoring and included in the LATITUDE database. Some were excluded: 37,570 who had their first LATITUDE transmission >3 months after pacemaker implantation, 26,976 who had <600,000 beats available, 26,963 who had device-determined AF (23,220: >1% of RA beats ≥170 beats/min and 3,743 subjects: mode switch episodes ≥5 minutes), 2,596 who had inconsistent programming in 3 months, and 8,332 who were programmed in a non-DDD/DDD-R mode. The remaining 34,543 subjects were followed a median of 2.8 (25^th^-75^th^, 1.0-4.0) years for the development of AF. Subject flowchart is shown in Figure 2.

**Figure 2.**
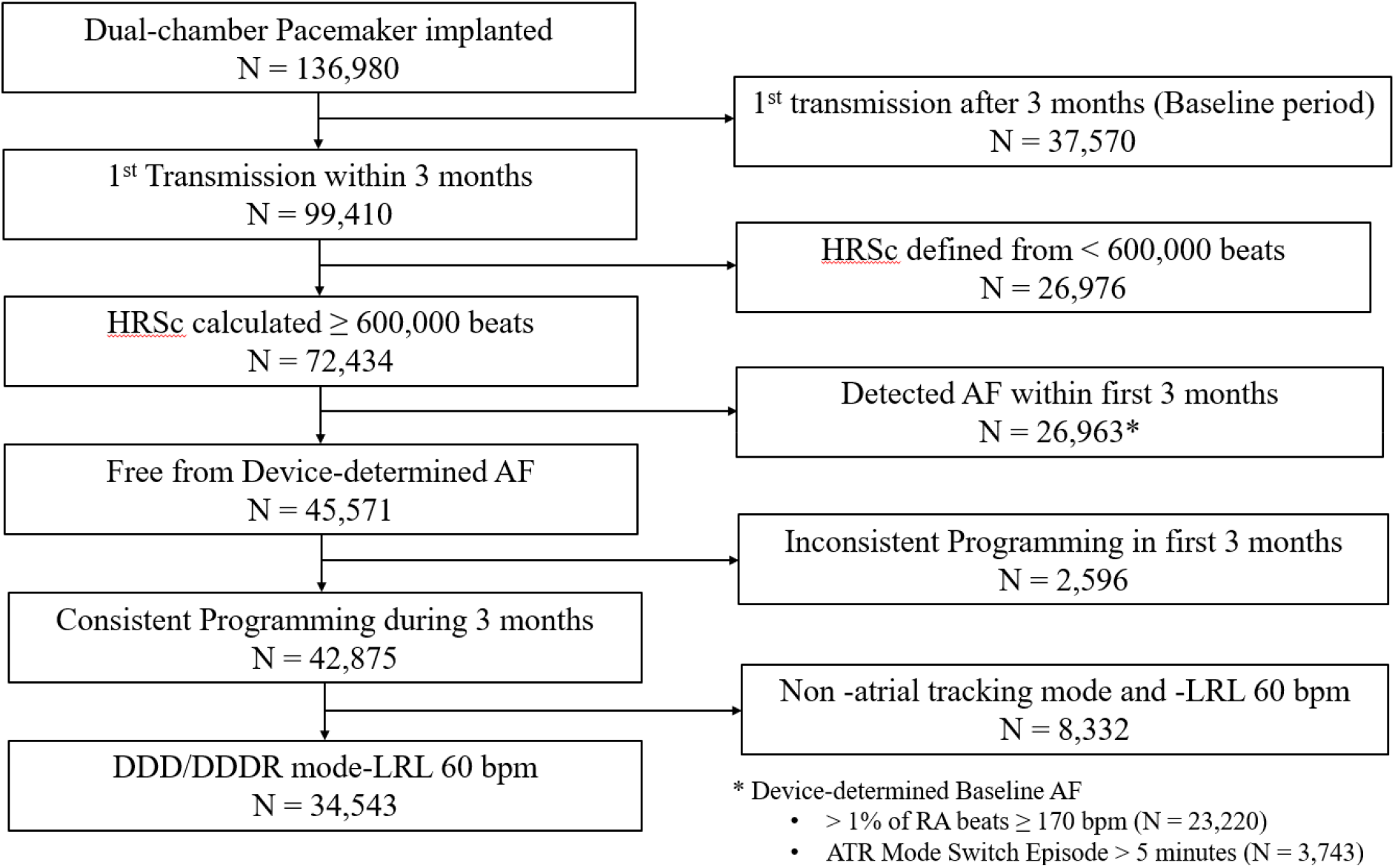
Patient Flowchart. This is the patient flowchart. A total of 136,980 dual-chamber pacemakers were implanted from 2013 to 2017 in subjects who were on remote monitoring and in the LATITUDE database. The following subjects were excluded: 1) Single chamber or biventricular pacemaker implantation, 2) First LATITUDE transmission >3 months after implantation, 3) HRSc calculated with <600,000 beats, 4) Device-determined AF detection during the first 3 months after implantation, 5) Crossover to another programming mode during the first 3 months after implantation, 6) Programming to non-atrial tracking mode (VVI, VVIR, DDI, DDIR) and lower pacing rate not equal to 60 bpm, Subjects from centers opting out of data sharing in a de-identified dataset for research were excluded. As a result, the remaining 34,543 subjects are the subjects for analysis. HRSc indicates heart rate score; AF, atrial fibrillation; RV, right ventricle; DDD, nonrate-responsive pacing; DDDR, rate-responsive pacing; LRL, lower rate limit.

Baseline demographics by HRSc grouping are shown in Table 1. Initial HRSc was significantly associated with age but not gender. Patients with initial HRSc≥70% were older, and had higher % RA pacing, lower % RV pacing and were more likely programmed with rate-response compared to subjects with initial HRSc <30-69% and <70%.

**Table 1.** Subject Demographics by HR Score Group.

| Characteristics | Statistic | HR Score, % |  |  | P-value |
| --- | --- | --- | --- | --- | --- |
|  |  | < 30<br>(N = 2,704) | 30-69<br>(N = 19,143) | ≥ 70<br>(N = 19,143) |  |
| Age at implantation, y | Mean ± SD | 69.6 ± 15.4 | 75.5 ± 11.8 | 78.4 ± 9.6 | < 0.001 |
| Females | N (%) | 1,311 (48) | 9,071 (47) | 5,919 (47) | 0.153 |
| Baseline % RA pacing, % | Median (IQR) | 4.7 (1.5, 9.5) | 15.8 (3.3, 32.8) | 77.6 (62.4, 90.9) | < 0.001 |
| Baseline % RV pacing, % | Median (IQR) | 49.8 (0.3, 99.3) | 27.5 (0.4, 99.1) | 3.3 (0.3, 80.6) | < 0.001 |
| Initial HR Score, % | Median (IQR) | 27 (25, 29) | 46 (38, 57) | 84 (78, 91) | < 0.001 |
| DDDR Mode | N (%) | 1,235 (46) | 11,137 (58) | 10,535 (83) | < 0.001 |

HR indicates heart rate; RA, right atrium; RV, right ventricle; DDDR mode, rate-responsive pacing mode; SD, standard deviation; IQR, interquartile range.

### Heart Rate Score

Initial HRSc was calculated at median of 57 days (interquartile range 37-77 days) after pacemaker implantation at 1^st^ LATITUDE transmission. The median value of the HRSc was 60% (interquartile range 40-82%). The group with <30% in initial HRSc accounted for 7.8% of total study subjects, the group with 30-69% initial HRSc accounted for 55.4%, and the group with ≥70% HRSc accounted for 36.8% (2,704 subjects, 19,143 subjects, 12,696 subjects of 34,543 total subjects).

### Primary End Point - Atrial Fibrillation

Over a median follow-up of 2.8 years (interquartile range: 1.0-4.0 years), 12,587 of 34,543 (37%) subjects (Kaplan-Meier rate of 54%) experienced the primary endpoint, >1% of atrial beats ≥170 beats/min through 7 years during post-baseline follow-up. Subjects with HRSc ≥70% at baseline, had a significantly increased risk of the primary endpoint definition of AF (log-rank *P* <0.001) during follow-up compared to subjects with HRSc <30% and subjects with HRSc between 30-69% (Figure 3). The incidence of AF increased in proportion to each 10% increase in HRSc (log-rank *P* <0.001).

**Figure 3.**
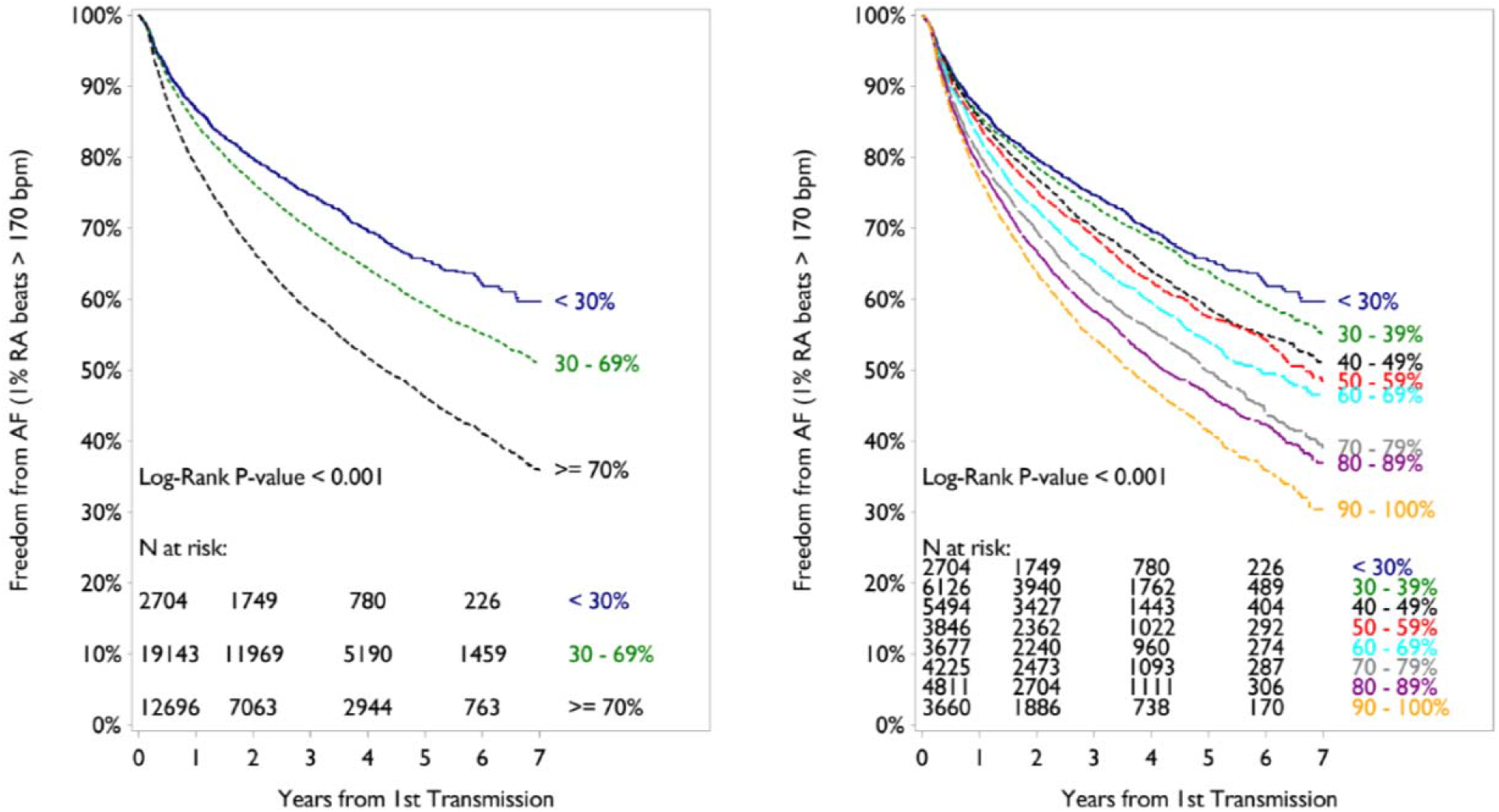
Survival free from AF by HRSc Group. Survival free from AF by HRSc Grouping is shown in this Kaplan-Meyer analysis. Left panel shows that subjects with ≥70% in initial HRSc had significantly increased risk of AF than subjects with <30% and 30-69% in initial HRSc, the primary end point. Right panel shows that the incidence of AF increased in proportion to every 10% HRSc. HRSc indicates heart rate score; AF, atrial fibrillation.

Multivariable analysis also showed that higher HRSc (HR 1.07, 95% CI 1.05-1.09; P < 0.0001) independently predicted subsequent occurrence of AF (based on the primary endpoint definition) after adjusting for age, gender, % RV pacing, and rate response programming (Table 2).

**Table 2.** Multivariable analysis to identify risk factors for AF including initial HR Score (per 10 percent) and possible predictors: age, gender, % RV pacing, rate response programming.

| Covariate | Hazard Ratio | Lower CI | Upper CI | Chi-Square |  |
| --- | --- | --- | --- | --- | --- |
|  |  |  |  | Stat | P-value |
| Baseline HR Score (per 10 %) | 1.067 | 1.045 | 1.089 | 37.6209 | < 0.0001 |
| Age (per 5 years) | 1.091 | 1.076 | 1.105 | 169.3069 | < 0.0001 |
| Female vs. Male | 0.972 | 0.926 | 1.020 | 1.3152 | 0.2515 |
| Baseline % RV Pacing (per 10%) | 0.991 | 0.985 | 0.997 | 8.6679 | 0.0032 |
| DDDR vs. DDD | 1.156 | 1.077 | 1.241 | 15.9470 | < 0.0001 |

HR indicates heart rate; RV, right ventricle; DDDR, rate-responsive pacing; DDD, nonrate-responsive pacing; CI, confidence interval.

#### Secondary Analyses

Considering each of the more restrictive definitions of AF (secondary endpoints: AF>5%, AF>10%, AF>25% of atrial beats ≥170 beats/min, and mode switch episodes ≥24 hours), subjects with initial HRSc ≥70% had significantly increased risk of subsequent AF versus those with initial HRSc <30% (*P* <0.001 by log-rank test) (Figure 4).

**Figure 4.**
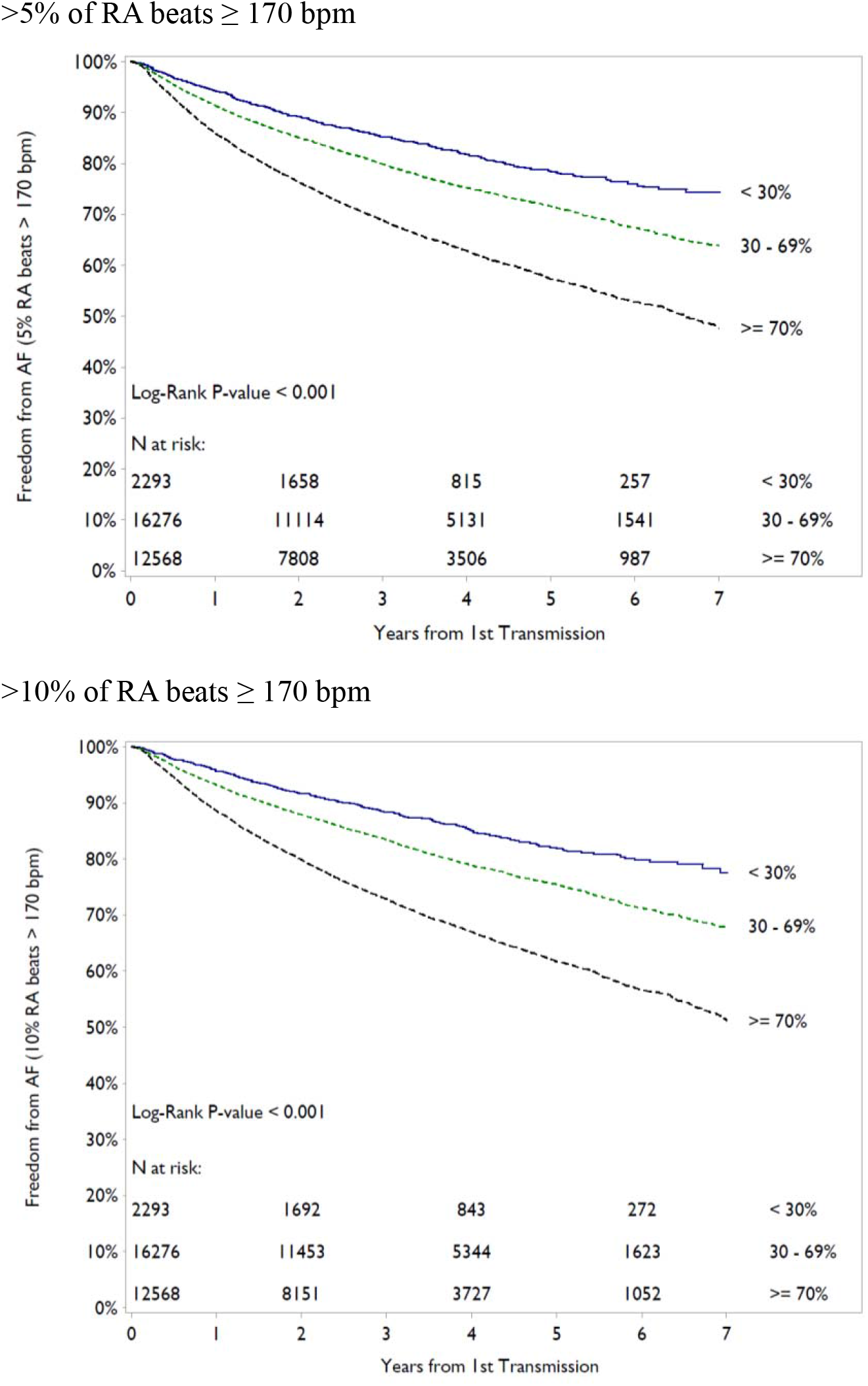
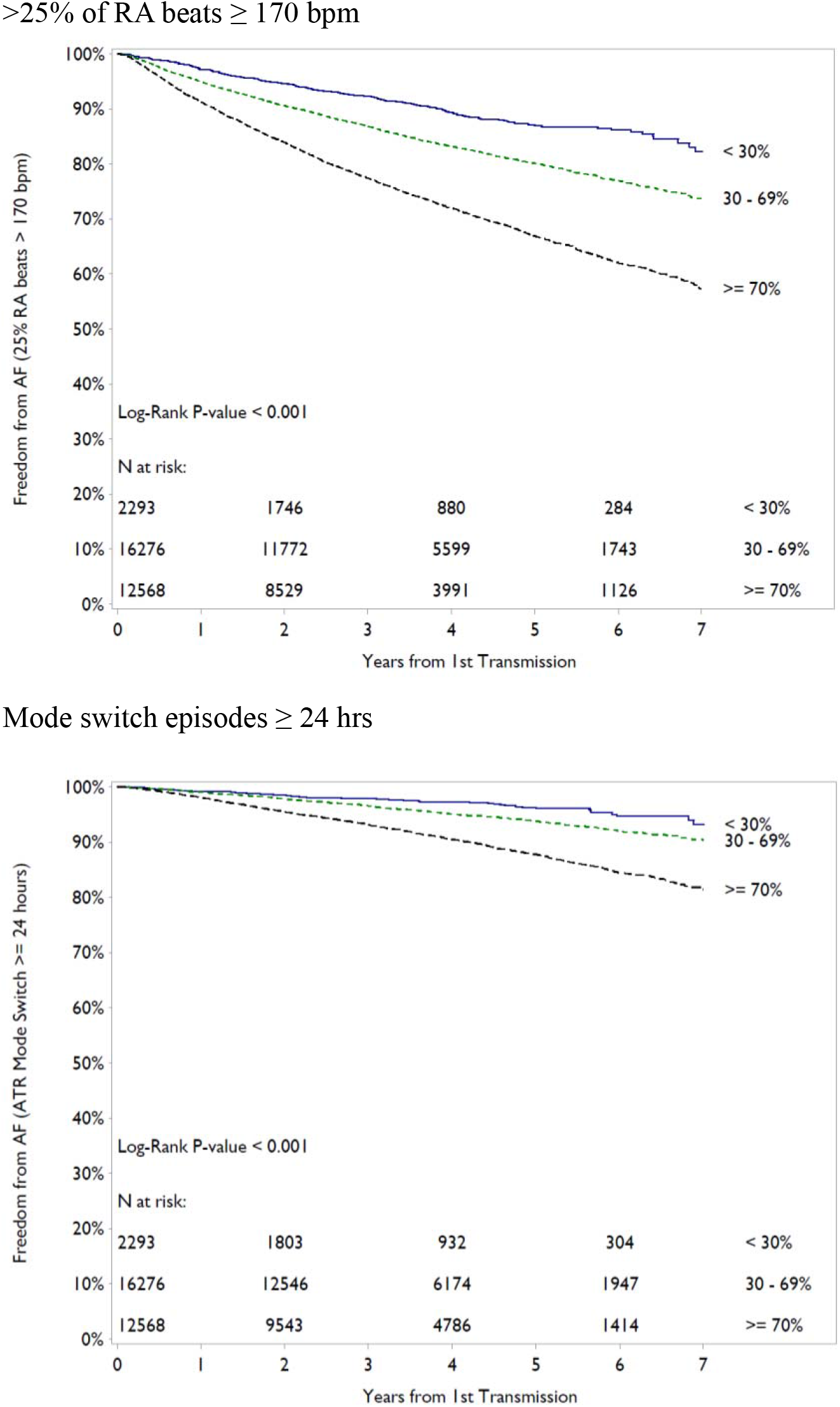
Results of Secondary Outcome. By each definition of device determined AF (>5%, >10% and >25% of atrial heart beats at more than 170bpm, and also with mode switch episodes lasting >24hrs), subjects with >70% in the initial HRSc had significantly increased risk of AF compared to subjects with <30% by log-rank test. HRSc indicates heart rate score; AF, atrial fibrillation; RA, right atrium.

### Association of Heart Rate Score and Percent Atrial Pacing and its relationship with the incidence of AF

Figure 5A shows a strongly positive correlation between %RA pacing during the baseline period and initial HRSc (Pearson correlation coefficient, r = 0.9). However in the patients with low %RA pacing (i.e., pacing ≤20%), the correlation was very weak (r = 0.13) (Figure 5B) and in the patients with RA pacing >20%, the correlation was much stronger (r = 0.84) (Figure 5C).

**Figure 5.**
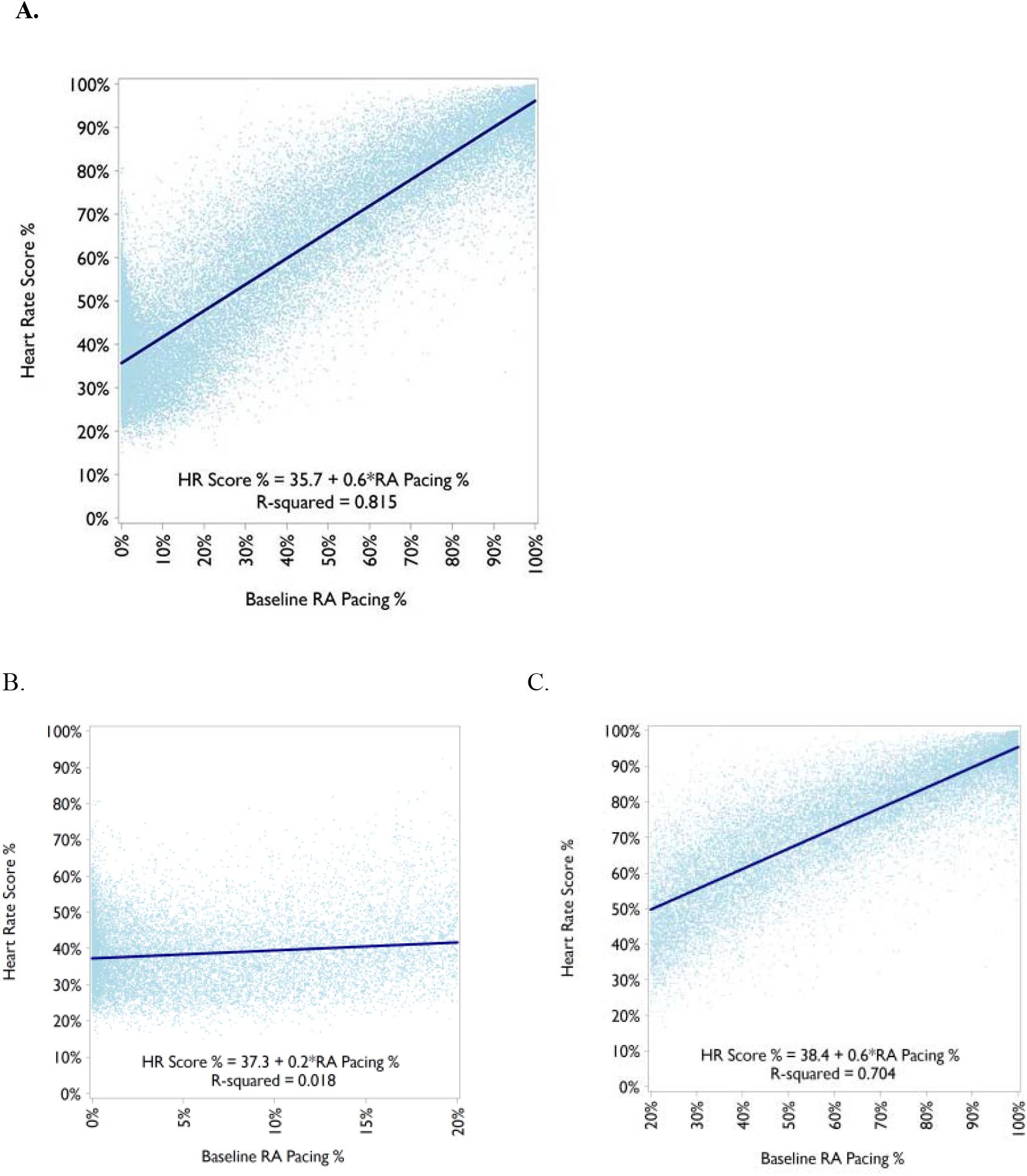
Heart Rate Score and RA pacing (Correlation by continuous measures) (A) The %RA pacing and initial HRSc were strongly correlated as a whole (Pearson correlation coefficient, r = 0.90). (B) In the patients in patients with RA pacing ≤20%, the correlation was very weak between the %RA pacing and initial HRSc (r = 0.13). (C) In the patients in patients with RA pacing >20%, the correlation was much stronger between the %RA pacing and initial HRSc (r = 0.84).

In both groups (RA pacing ≤20% and >20%), subjects with an initial HRSc ≥70% had a significantly higher incidence of AF versus those with an initial HRSc <70% by log-rank test (*P* <0.001) (Figure 6).

**Figure 6.**
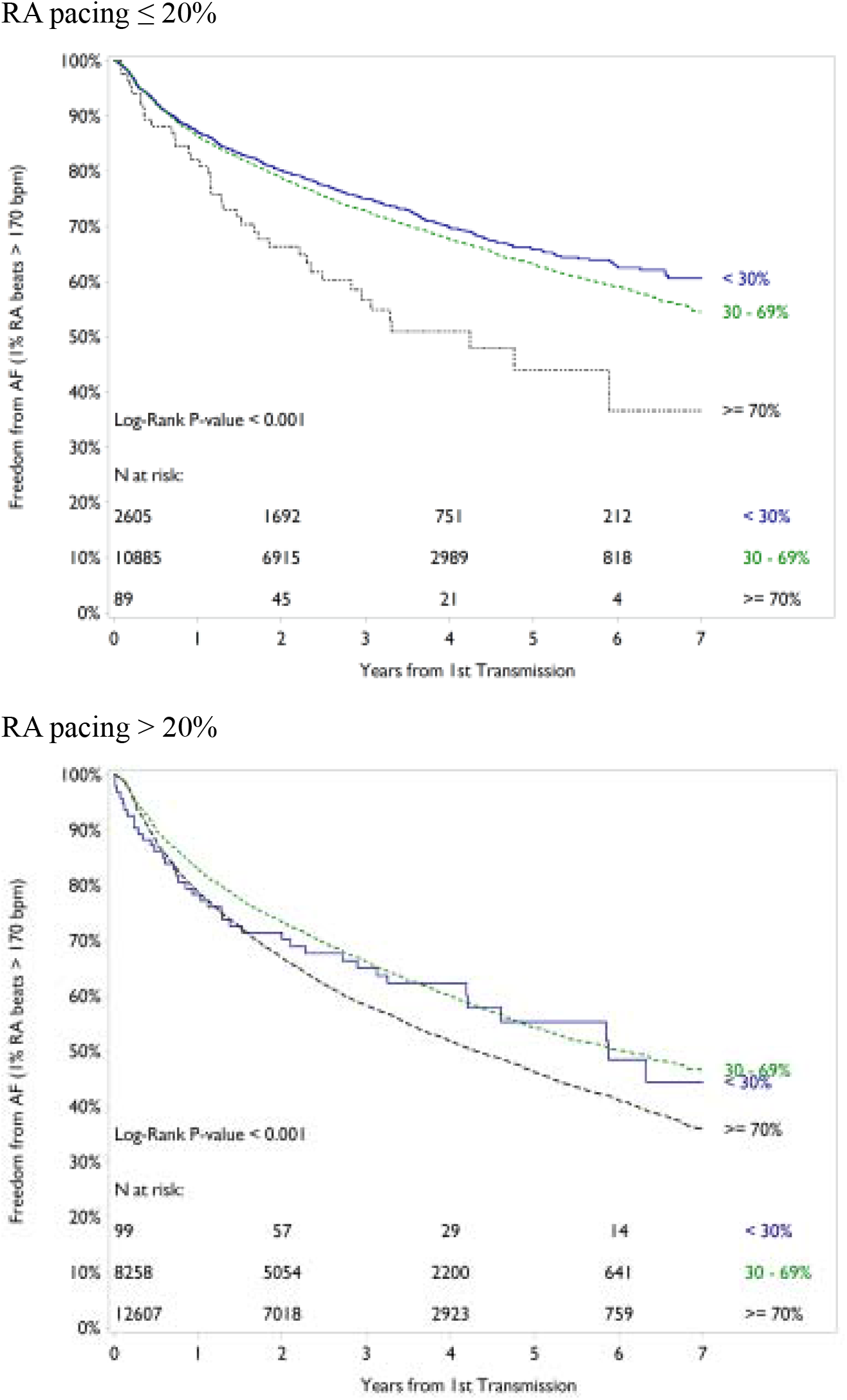
Free from AF by Heart Rate Score in Patients with Low and High RA pacing. In both groups (RA pacing ≤ 20% and > 20%), subjects with ≥70% in the initial HRSc had significantly increased risk of AF compared to the subjects with < 70% by log-rank test.

## Discussion

### Main Findings

The main finding of this study is that initial HRSc, calculated within 3 months after dual-chamber PM implantation, independently predicted subsequent new-onset device defined AF. Subjects with initial HRSc ≥70% had an associated increased risk of AF by all device determined metrics. This analysis held true over a 2.8 year follow-up, after adjustment for risk factors for AF and with various definitions of AF (1%, >5%, >10%, >25% of atrial beats ≥170 beats/min and mode switch episodes ≥24hrs). Furthermore, a higher greater initial HRSc was an independent multivariable predictor of subsequent AF. Despite the correlation between %RA pacing and initial HRSc, the prediction of AF development was still significant when there was low %RA pacing. These data are the first to show a relationship between HRSc and new-onset AF in a pacemaker population.

### Association between Heart Rate Score and Incidence of Atrial Fibrillation

HRSc is a metric of chronotropic incompetence in PM patients.^7^ Patients with little or no spontaneous heart rate variation above the lower rate limit (LRL) have a high HRSc because most atrial events occur at the programmed LRL. Chronotropic incompetence typically occurs in patients with concomitant sinus node disease and many patients with sinus node disease are well known to develop subsequent AF.^8,9^ These facts raised the possibility that patients with more severe chronotropic incompetence and thus higher HRSc are susceptible to developing AF.

O’Neal reported that patients with inadequate heart rate response during exercise are at increased risk to develop AF by using maximal predicted heart rate values <85% and chronotropic index values <80% during exercise-treadmill stress testing defined as chronotropic incompetence.^10^ Chronotropic incompetence may also be associated with heart failure,^11-13^ and heart failure may in turn be related to AF.^14^ Accordingly, the association between higher HRSc and AF may be related to chronotropic incompetence but alternatively, relatively inadequate heart rate for the physiologic demand may also contribute to AF.^10^

### Heart Rate Score – A Novel Predictor of Atrial Fibrillation in Pacemaker Patients

Although several risk factors for AF have been reported, little established electrocardiographic or pacemaker programmed or measured metrics have been determined to be predictors of AF. Associations between %RA pacing and AF have been identified. A meta-analysis including of 1,507 patients from four pacemaker studies showed that atrial pacing was associated with a higher incidence of AF in patients with sinus node disease.^15^ However, a sub-analysis from Danish multicenter randomized on AAI versus DDD pacing in sick sinus syndrome (DANPACE) trial with 396 patients showed that no association was identified between %RA pacing and AF in patients with sinus node disease.^16^ In addition, a positive correlation between the percent ventricular pacing and incident AF has also been reported.^17-19^ Finally, the paced P-wave duration was shown to predict AHREs in patients with implantable cardiac devices.^20^ This study highlights that the baseline HRSc can predict AF in pacemaker patients independent of well-known risk factors.

### The Clinical Importance of Atrial Fibrillation Detected by Cardiac Device

It is noteworthy that the results were robust and replicated with multiple metrics used to define AF not only by AF burden (1%, >5%, >10%, >25%) but also by ATR mode switch episodes >24 hours. When AF burden was measured by the most sensitive metric, >1% AF burden (primary outcome), this metric may be influenced by frequent premature atrial contractions. Several studies report that the burden of atrial ectopy is associated independently with new-onset AF, pacemaker implantation, cardiovascular hospitalization, and long term mortality.^21-24^

Subjects with an initial HRSc ≥70% had an associated increased risk of AF and when AF is defined by ATR mode switch duration >24 hours, it is more likely to represent persistent AF. A sub-analysis of the ASSERT trial showed that patients with AHREs >24 hours had significantly higher subsequent risk of subsequent stroke or systemic embolism (adjusted hazard ratio 3.24, 95% CI 1.51-6.95) versus those with no AHREs.^6^ Another sub-analysis of ASSERT trial showed that for patients with cardiac devices, AHREs >24 hours was strongly associated with heart failure hospitalization.^25^

### Relationship between Heart Rate Score and Percent Right Atrial Pacing

There was a strong correlation between %RA pacing and initial HRSc when all patients were considered (figure 5). For patients with a low %RA pacing, this correlation was weak but in the patients with high %RA pacing, the correlation was stronger. Thus, HRSc and %RA pacing do not necessarily indicate the same relationship; HRSc can predict AF even when the %RA pacing is low.

The LRL setting affects both %RA pacing and HRSc. Programming a high LRL can override the resting intrinsic rate and move atrial events to the lowest heart rate bin (including the LRL). This results in an increase in %RA pacing and increase in HRSc. Conversely, programming a lower LRL can result in a decreased %RA pacing and lower HRSc. As we focused on patients set at an LRL 60 beats/min, we could not assess the impact of different LRL settings. Further research is needed to assess the relationship of LRL on AF, %RA pacing and HRSc.

### Study Limitations

This study is a retrospective analysis and, therefore, hypothesis generating. Prospective studies would be useful if achievable with sufficient sample size, but the sample size in this investigation allowed for analysis untenable in a smaller study. The database did not collect all clinical information and potential prognosticators. The present study also does not answer whether lowering HRSc reduces new-onset AF. Finally we cannot determine if HRSc is completely independent of the %RA pacing.

## Conclusion

The initial HRSc in patients undergoing pacemaker implant predicts new-onset, device-determined, AF regardless the definition of AF utilized.

## Data Availability

Data and the datasets during and/or analyzed during the current study are available from the corresponding author on reasonable request.

## Notes

### Competing Interest Statement

The authors have declared no competing interest.

### Clinical Trial

Current study is retrospective study.

### Funding Statement

No external funding was receieved.

### Author Declarations

This study, approved by the Boston Scientific Governance Board (Boston Scientific Company, Boston, USA), is a retrospective analysis of deidentified data from the LATITUDE remote monitoring cardiac implantable electronic device database.

